# Chikungunya Virus VLP Vaccine: Phase 3 Trial in Adolescents and Adults

**DOI:** 10.1101/2024.10.11.24315179

**Authors:** Jason S. Richardson, Deborah M. Anderson, Jason Mendy, Lauren C. Tindale, Sufia Muhammad, Tobi Loreth, Sarah Royalty Tredo, Kelly L. Warfield, Roshan Ramanathan, Jorge T. Caso, Victoria A. Jenkins, Patrick Ajiboye, Lisa Bedell, the EBSI-CV-317-004 Study Group

## Abstract

1

**Background:** Chikungunya disease is a growing global public health concern. Chikungunya virus (CHIKV) virus-like particle (VLP) vaccine is a single dose, pre-filled syringe for intramuscular injection.

**Methods:** A pivotal phase 3 randomized, double-blind, placebo-controlled trial was conducted in healthy adolescents and adults 12-64 years of age. Participants received either CHIKV VLP vaccine or placebo on Day 1. Immunogenicity objectives assessed CHIKV serum neutralizing antibody (SNA) titers (NT_80_) at selected timepoints. Safety was assessed through Day 183 postvaccination.

**Results:** A total of 2790 participants received CHIKV VLP vaccine and 464 received placebo (n=3454). Coprimary endpoints were met including 1) immunologic superiority to placebo and 2) geometric mean titer at Day 22. Results demonstrated that CHIKV VLP vaccine induced a protective seroresponse (SNA NT_80_ ≥100; considered the presumptive seroprotective antibody response), in 97% of individuals at Day 15 (2 weeks), in 98% of individuals at Day 22, and in 86% of individuals at Day 183 (6 months). CHIKV VLP vaccine demonstrated a favorable safety profile. The majority of adverse events were self-limited and grade 1 or 2 in severity. The most common adverse events were injection site pain, fatigue, headache, and myalgia.

**Conclusions:** CHIKV VLP vaccine induces a rapid and robust immune response by Day 15 that lasts through Day 183 postvaccination. These findings support the potential for this vaccine to protect individuals 12-64 years of age from disease caused by CHIKV. (Funded by Emergent BioSolutions Inc. and Bavarian Nordic A/S [as successor in interest to Emergent BioSolutions Inc.]; ClinicalTrials.gov number, NCT05072080).

## 2 INTRODUCTION

Chikungunya disease, caused by chikungunya virus (CHIKV), is transmitted to humans through the bite of infected mosquitoes. The classic symptom of acute CHIKV disease is a debilitating polyarthralgia that is present in greater than 90% of cases.^1^ Other acute clinical manifestations of infection include high fever, rash, headache, nausea, fatigue, and myalgia.^1,2,3,4^ Acute symptoms may persist for 1 to 2 weeks after infection and can significantly impact daily productivity, primarily due to the reduced mobility associated with joint pain. In some cases, chronic symptoms persist for months or years with a critical impact on quality of life.^5,6^ Currently, one live-attenuated vaccine is licensed for the prevention of disease caused by CHIKV in individuals 18 years of age and older who are at increased risk of exposure to CHIKV.^7,8^ There is no marketed vaccine available for adolescents under the age of 18.

CHIKV virus-like particle (VLP) vaccine (previously PXVX0317) comprises the E1, E2, and C proteins of CHIKV Senegal West African strain 37997 that are assembled to form an approximately spherical, highly ordered VLP.^9^ Notably, the E1 glycoprotein mediates membrane fusion during virus infection of host cells and E2 transmembrane glycoprotein is responsible for receptor binding to host cells during viral replication,^10^ and has been shown to be the primary target of CHIKV neutralizing antibodies induced during natural infection.^11^ Protection against subsequent infection has been shown to correlate with the presence of CHIKV antibodies that neutralize the virus *in vitro*.^12^ CHIKV VLP vaccine is comprised of 40 μg CHIKV VLP in buffered solution and adjuvanted with aluminum hydroxide. The product is supplied as a pre-filled syringe for ease of use and increased dosing accuracy. Here we report safety, tolerability, and immunogenicity data for CHIKV VLP vaccine in healthy adolescents and adults 12 to 64 years of age.

## 3 METHODS

### 3.1 Trial design, oversight, and participants

This pivotal phase 3, randomized, placebo-controlled, double-blind, parallel group trial was conducted at 47 sites in the US. Eligible participants were healthy adolescents and adults 12 to 64 years of age. Full inclusion and exclusion criteria are provided in the Supplementary Appendix. The trial was conducted in compliance with International Council on Harmonization Good Clinical Practice guidelines and the principles in the Declaration of Helsinki. The trial protocol was approved by an independent institutional review board and by federal regulatory agencies. Written informed consent and/or assent was obtained prior to any trial procedure. An independent Safety Monitoring Committee reviewed blinded safety data at two timepoints: 1) after the first 300 participants completed the Day 29 visit and 2) after all participants completed the Day 29 visit. Bavarian Nordic A/S and Emergent BioSolutions Inc. were responsible for trial design, data collection, data analysis, data interpretation, and writing of the report. Additional details regarding the design of the trial are provided in the protocol, available at NEJM.org.

### 3.2 Randomisation and treatment

Participants were randomized in a 2:2:2:1 ratio within each of three age groups (12 to 17, 18 to 45, and 46 to 64 years of age) at each site to receive one of 3 consecutively manufactured lots of CHIKV VLP vaccine or placebo. Participants attended a screening visit, then a Day 1 visit which included randomization, blood collection, and administration of a single dose of CHIKV VLP vaccine or placebo by intramuscular injection in the deltoid muscle (Figure S1). Day 8, 15, 22, and 183 visits included review of adverse events (AEs) and concomitant medications and blood collection. Day 29 and 92 phone visits included review of AEs.

### 3.3 Immunogenicity

A luciferase-based CHIKV human SNA assay was used to measure 80% serum neutralizing antibody (SNA) titer (NT_80_) against CHIKV in human serum samples (Supplementary Appendix). Seroresponse rate (SRR) was defined as the percentage of participants who achieved SNA NT_80_ ≥100 (presumptive seroprotection rate agreed with the FDA and EMA).

The coprimary endpoints were: 1) difference in CHIKV SNA SRR (vaccine minus placebo) at Day 22, 2) CHIKV SNA geometric mean titer (GMT) at Day 22 for vaccine and placebo, and 3) CHIKV SNA GMT ratio at Day 22 between all 3 pairs of vaccine lots (A:B, B:C, A:C) in adults 18 to 45 years of age.

The key secondary endpoints were the difference in CHIKV SNA SRR at Day 15, Day 183, and Day 8, in that order. Other secondary immunogenicity endpoints included 1) CHIKV SNA GMTs by trial arm at Day 8, Day 15, and Day 183, 2) geometric mean fold increase (GMFI) from Day 1 to subsequent collection timepoints, and 3) number and percentage of participants with SNA NT_80_ ≥15, and 4-fold rise over baseline.

### 3.4 Safety

Participants were monitored for signs of an acute adverse reaction(s) to vaccination for 30 minutes. Solicited AEs were collected from injection administration on Day 1 through Day 8 using a diary. Solicited AEs included local events of pain, redness, and swelling at the injection site and systemic events of oral temperature ≥38.0°C (≥100.4°F), chills, fatigue, headache, myalgia, arthralgia, and nausea. Unsolicited AEs were collected from Day 1 through Day 29. Serious AEs (SAEs), AEs of special interest (AESI) (defined as new onset or worsening arthralgia that was medically attended), and medically attended AEs (MAAEs) were collected from Day 1 postvaccination through Day 183. The investigator graded all AEs for severity as grade 1 (mild), grade 2 (moderate), grade 3 (severe), or grade 4 (potentially life-threatening). The investigator assessed AE causality. If the relationship between the AE and the trial drug was determined to be “possible” or “probable,” the event was considered “related.”

### 3.5 Statistical analysis

The sample size for the trial was partially based on the regulatory need for a specific number of participants treated with CHIKV VLP vaccine for safety. With an assumed 10% rate of nonevaluable participants and the difference between CHIKV VLP vaccine and placebo assumed to be approximately 85-90%, the power to show superiority over placebo with 2430 CHIKV VLP vaccine and 405 placebo evaluable participants was >99.9% for the combined age groups. The difference in SRR between vaccine and placebo groups that was considered clinically relevant was 70%. With 2430 vaccine-treated participants and a target SRR of 90% vs a rate of 0% for placebo, the width of a 2-sided 95% CI would be ±1.2%. The difference in SRR had to be above 72% for the lower bound of the 95% CI for the difference to be ≥70%.

The exposed population included all participants who received a CHIKV VLP vaccine or placebo injection. The safety population included all participants from the exposed population who provided safety assessment data. The modified intent-to-treat (mITT) population included all participants who were vaccinated and had at least one post-injection CHIKV SNA titer result. The immunogenicity evaluable population (IEP) included mITT population participants who had no measurable CHIKV SNA at Day 1 (below lower limit of quantitation [LLOQ]), had evaluable CHIKV SNA at Day 22, and had no reason for exclusion as defined prior to unblinding (primary population for immunogenicity analysis). Additional details are provided in the statistical analysis plan, available at NEJM.org.

The primary objectives of the trial were based on SNA NT_80_ titers calculated to determine the CHIKV neutralizing antibody response. The difference between treatment groups in SRR (proportion of participants with SNA NT_80_ ≥100) was tested using a chi-square test with a 2-sided alpha=0.05 in the IEP across all age groups combined and the 2-sided 95% CI for the difference was computed using the Newcombe hybrid score method. In the computation of the GMTs, values below the assay LLOQ of <15 were assigned the value LLOQ/2=7.5. The GMTs were compared between CHIKV VLP vaccine and placebo treatment groups and were analyzed via a linear model based on a two-sided alpha=0.05. The primary model was an ANOVA, with logarithmically transformed CHIKV SNA titers (log_10_) as the dependent variable and treatment group and trial site as the fixed effects in the model. The adjusted least square means and their 95% CIs calculated based on the ANOVA were back transformed and reported as the group GMT values. The proportion of participants with seropositivity (SNA NT_80_ ≥15; above the LLOQ) and those with a 4-fold increase in titer over baseline were also summarized. The lot consistency was assessed using a similar ANOVA to the GMT but using only the CHIKV VLP vaccinated participants and vaccine lot, rather than vaccine group, as a factor in the model. Multiplicity was addressed by requiring both coprimary endpoints to be met as a success criterion and via hierarchical testing for the key secondary endpoints.

Safety data, frequency and percentage of participants with either solicited or unsolicited AEs, were summarized descriptively including duration, severity and causality of the events observed.

## 4 RESULTS

### 4.1 Participants

A total of 4215 participants were screened for this trial between September 29, 2021, and September 23, 2022, of which 3258 were eligible and enrolled, and 3254 (99.9%) received investigational product. The IEP included 2983 participants, of which 2559 received CHIKV VLP vaccine (841, 860, 858 received lot A, B, and C, respectively) and 424 received placebo (**Figure 1**).

**Figure 1:**
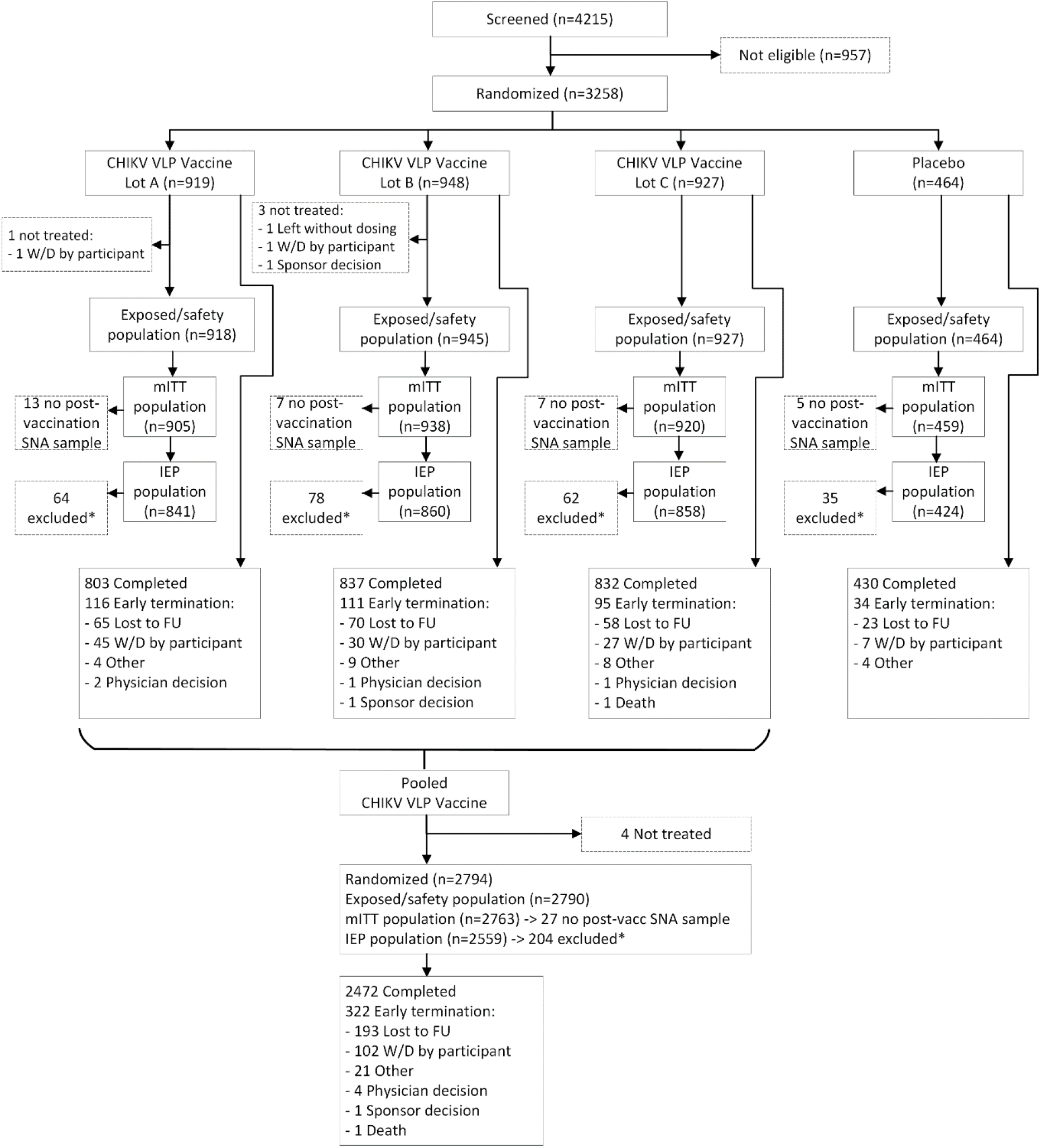
Screening, Enrollment, and Randomization. *Exclusions from IEP were due to: no SNA result in Day 22 window, measurable Day 1 SNA (baseline positive), prohibited vaccine/medication use, eligibility criteria not met, missing Day 1 SNA sample, and/or serum processing error. FU = follow up; IEP = immunogenicity evaluable population; LLOQ = lower limit of quantitation; mITT = modified intent-to treat; SNA = serum neutralizing antibody; W/D = withdrawn.

There was no appreciable difference between the CHIKV VLP vaccine and placebo groups in demographic characteristics (**Table 1**). In the randomized population, 48.8% of participants were male. There were 254 adolescent participants aged 12 to 17 years (217 vaccine, 37 placebo), 1906 participants aged 18 to 45 years (1636 vaccine, 270 placebo), and 1098 participants aged 46 to 64 years (941 vaccine, 157 placebo). White participants accounted for 73.2% of the randomized and 19.1% were Black or African American.

**Table 1:** Baseline Characteristics of the Randomized Population.

|  | **CHIKV VLP Vaccine**<br>**(N=2794)** | **Placebo**<br>**(N=464)** | **Total**<br>**(N=3258)** |
| --- | --- | --- | --- |
| **Age (years)** |  |  |  |
| Mean (SD) | 39 (14.3) | 39 (14.4) | 39 (14.3) |
| Median | 38 | 38 | 38 |
| Min, Max | 12, 64 | 12, 64 | 12, 64 |
| **Sex – n (%)** |  |  |  |
| Male | 1358 (48.6) | 233 (50.2) | 1591 (48.8) |
| Female | 1436 (51.4) | 231 (49.8) | 1667 (51.2) |
| Females of childbearing potential | 846 (58.9) | 136 (58.9) | 982 (58.9) |
| **Age group – n (%)** |  |  |  |
| 12 to 17 years | 217 (7.8) | 37 (8.0) | 254 (7.8) |
| 18 to 45 years | 1636 (58.6) | 270 (58.2) | 1906 (58.5) |
| 46 to 64 years | 941 (33.7) | 157 (33.8) | 1098 (33.7) |
| **Race – n (%)** |  |  |  |
| White | 2043 (73.1) | 341 (73.5) | 2384 (73.2) |
| American Indian or Alaska Native | 30 (1.1) | 2 (0.4) | 32 (1.0) |
| Asian | 79 (2.8) | 16 (3.4) | 95 (2.9) |
| Black or African American | 534 (19.1) | 89 (19.2) | 623 (19.1) |
| Native Hawaiian or Other Pacific Islander | 6 (0.2) | 4 (0.9) | 10 (0.3) |
| Multiracial | 78 (2.8) | 8 (1.7) | 86 (2.6) |
| Not reported | 24 (0.9) | 4 (0.9) | 28 (0.9) |
| **Ethnicity – n (%)** |  |  |  |
| Hispanic or Latino | 506 (18.1) | 71 (15.3) | 577 (17.7) |
| Not Hispanic or Latino | 2226 (79.7) | 379 (81.7) | 2605 (80.0) |
| Not reported | 61 (2.2) | 14 (3.0) | 75 (2.3) |
| Unknown | 1 (<0.1) | 0 | 1 (<0.1) |
| **Height (cm)** |  |  |  |
| Mean (SD) | 170.8 (9.9) | 171.2 (10.0) | 170.9 (9.9) |
| Median | 170.2 | 170.8 | 170.2 |
| Min, Max | 133.4, 203.2 | 141.2, 201.2 | 133.4, 203.2 |
| **Weight (kg)** |  |  |  |
| Mean (SD) | 78.4 (16.3) | 77.6 (16.4) | 78.3 (16.3) |
| Median | 77.8 | 76.0 | 77.5 |
| Min, Max | 30.8, 132.6 | 35.8, 128.2 | 30.8, 132.6 |
| **BMI (kg/m<sup>2</sup>)** |  |  |  |
| Mean (SD) | 26.8 (4.5) | 26.4 (4.6) | 26.7 (4.5) |
| Median | 26.8 | 26.3 | 26.7 |
| Min, Max | 13.1, 35.4 | 15.7, 34.9 | 13.1, 35.4 |
| **Baseline CHIKV SNA Serostatus – n (%)** |  |  |  |
| Negative (<LLOQ) | 2725 (97.5) | 458 (98.7) | 3183 (97.7) |
| Positive (≥LLOQ) | 63 (2.3) | 6 (1.3) | 69 (2.1) |
| Missing | 6 (0.2) | 0 | 6 (0.2) |

BMI = body mass index; CHIKV VLP = chikungunya virus virus-like particle; LLOQ = lower limit of quantitation; Min = minimum; Max = maximum; N = total number of participants in the group; n = number of participants per parameter; SD = standard deviation; SNA = serum neutralizing antibody.

### 4.2 Immunogenicity

At Day 22, 97.8% (2503/2559) of participants in the CHIKV VLP vaccine group had a seroresponse (SNA NT_80_ ≥100), compared to 1.2% (5/424) of participants in the placebo group for the IEP (**Table 2**). The SRR difference was 96.6% (95% CI: 95.0%, 97.5%) which was statistically significant (P<0.0001, chi-square test) and clinically relevant (lower bound of the 95% CI ≥70%). CHIKV SNA GMT at Day 22 for the vaccine group was 1618, while the placebo group was 7.9 for the IEP (Table S1 in the Supplementary Appendix); the vaccine group GMT was significantly higher than that for placebo (P<0.0001, ANOVA). Additionally, at Day 22, the pairwise lot comparison of SNA response to CHIKV VLP vaccine in adults aged 18 to 45 years demonstrated equivalence, so all coprimary endpoints were successfully achieved.

**Table 2.** CHIKV Serum Neutralizing Antibody Seroresponse Rate and Seroresponse Rate Difference by Trial Visit (Immune Evaluable Population).

| Day | Parameter | Seroresponse Rate<br>CHIKV VLP Vaccine<br>(N=2559) | Seroresponse Rate<br>Placebo<br>(N=424) | Seroresponse Rate<br>Difference | P value |
| --- | --- | --- | --- | --- | --- |
| Day 8 | n/N (%) | 1169/2510 (46.6%) | 2/419 (0.5%) | 46.1% | <0.0001 |
|  | 95% CI | 44.6%, 48.5% | 0.1%, 1.7% | 43.8%, 48.1% |  |
| Day 15 | n/N (%) | 2355/2434 (96.8%) | 3/395 (0.8%) | 96.0% | <0.0001 |
|  | 95% CI | 96.0%, 97.4% | 0.3%, 2.2% | 94.3%, 96.8% |  |
| Day 22 | n/N (%) | 2503/2559 (97.8%) | 5/424 (1.2%) | 96.6% | <0.0001 |
|  | 95% CI | 97.2%, 98.3% | 0.5%, 2.7% | 95.0%, 97.5% |  |
| Day 183 | n/N (%) | 1967/2301 (85.5%) | 6/401 (1.5%) | 84.0% | <0.0001 |
|  | 95% CI | 84.0%, 86.9% | 0.7%, 3.2% | 81.7%, 85.6% |  |

Seroresponse rate difference is (vaccine minus placebo); 95% CIs are based on the Newcombe hybrid score method. P value is from a two-sided chi-square test of equality of seroresponse percentages between groups. 95% CIs of seroresponse rates are based on the Wilson method. CHIKV VLP = chikungunya virus virus-like particle; CI = confidence interval; IEP = immunogenicity evaluable population; n = number of participants with seroresponse (SNA titer $\geq 100$ ); N = total number of participants in the group; SNA = serum neutralizing antibody; N/A = not applicable.

At Day 15, 96.8% of participants in the CHIKV VLP vaccine group had a seroresponse, compared to 0.8% in the placebo group (**Table 2**). The SRR difference at Day 15 was 96.0% [95% CI: 94.3%, 96.8%], which was statistically significant (P<0.0001, chi-square test) and clinically relevant. At Day 183, 85.5% of participants in the vaccine group had a seroresponse, compared to 1.5% in the placebo group (P<0.0001, chi-square test). At Day 8, 46.6% of participants in the vaccine group had a seroresponse, compared to 0.5% in the placebo group (P<0.0001, chi-square test).

Analysis by age group showed that the younger age groups elicited a more rapid and longer lasting response (**Figure 2A**, Table S2 in the Supplementary Appendix). While the SRR was relatively similar with a peak in all age subgroups at Day 15 and Day 22, the largest differences were seen at Day 8 and Day 183. GMT in the vaccine group was highest in the adolescent 12 to 17-year-old group (164 at Day 8, 1706 at Day 15, 2502 at Day 22, 511 at Day 183), followed by the 18 to 45-year-old group (115 at Day 8, 1369 at Day 15, 1878 at Day 22, 323 at Day 183), and then the 46 to 64-year-old subgroup (60 at Day 8, 709 at Day 15, 1175 at Day 22, 322 at Day 183) (**Figure 2B**).

**Figure 2.**
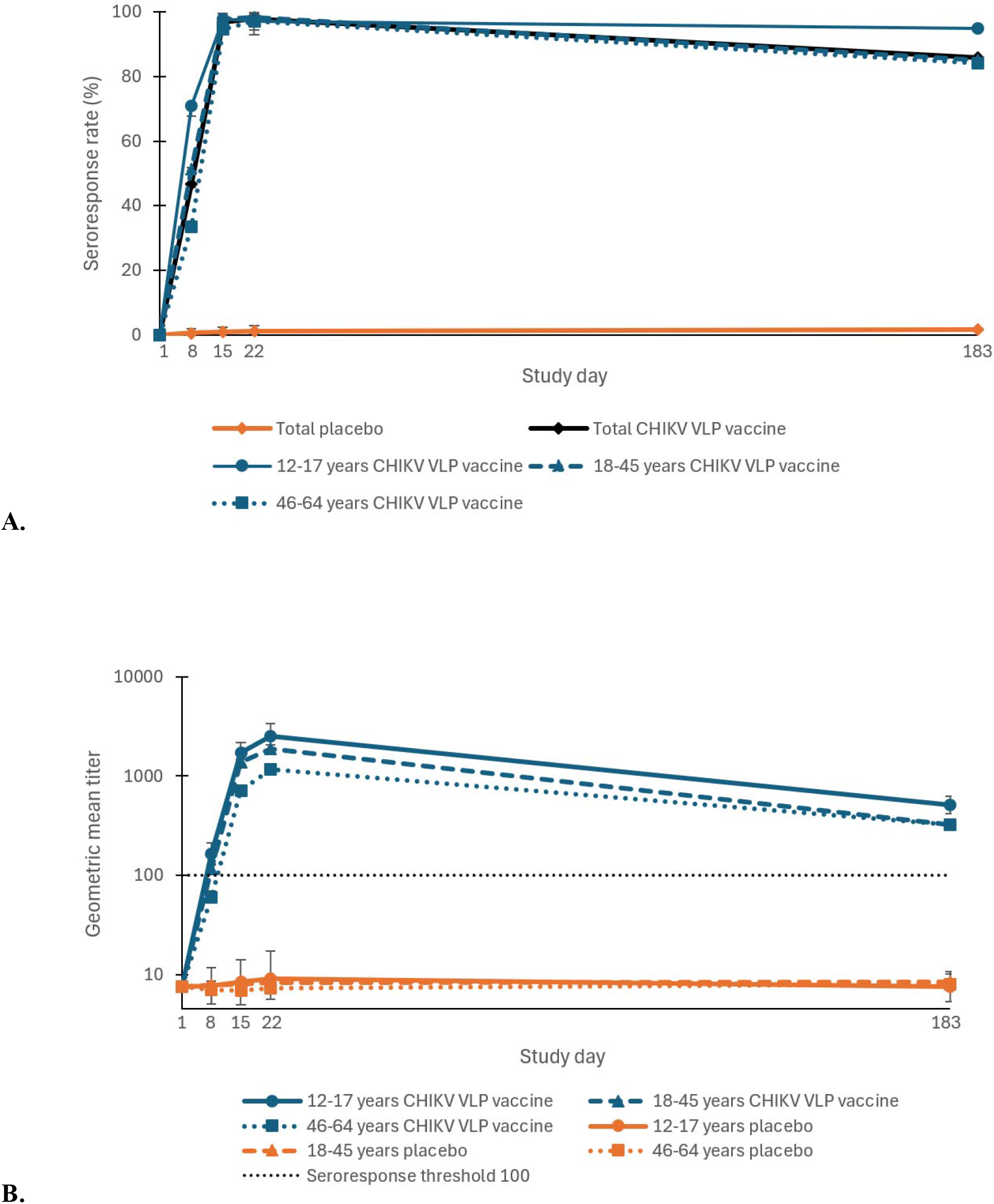
A. Seroresponse Rate and B. Geometric Mean Titers for CHIKV Serum Neutralizing Antibodies by Visit and by Age Group (Immunogenicity Evaluable Population). Y-axis is represented on a log10 scale. Vertical bars denote the 95% confidence intervals. Values below lower limit of quantitation (LLOQ=15) were assigned the value LLOQ/2=7.5.

CHIKV VLP vaccine had high seroconversion (SNA NT_80_ ≥15) and 4-fold rise over baseline titers at all timepoints (Table S3 in the Supplementary Appendix). Seroconversion was seen in 91.9% of vaccine recipients at Day 8, 99.5% at Day 15, 99.2% at Day 22, and 99.0% at Day 183. Placebo group participants who had a seroresponse at any timepoint (n=16) were investigated and identified as potential sample handling errors, as each of these participants had SNA NT_80_ ≥100 result at a single timepoint only, with SNA NT_80_ <15 at all other sample collection times.

A post ad hoc analysis found that baseline seropositive (defined as Day 1 predose SNA NT_80_ ≥15) CHIKV VLP vaccine group participants’ (n=63) immune response could be boosted by the CHIKV VLP vaccine and tended to have higher SNA titers postvaccination than that for baseline seronegative vaccine group participants. Baseline seropositive vaccine recipients had a Day 1 (baseline) GMT of 253, Day 8 GMT of 944, Day 15 GMT of 2878, Day 22 GMT of 3645, and Day 183 GMT of 862. Comparatively, baseline seronegative vaccine recipients (the IEP) had a Day 1 GMT below the LLOQ, Day 8 GMT of 93, Day 15 GMT of 1096, Day 22 GMT of 1618, and Day 183 GMT of 338. Additionally, baseline seropositive placebo participants’ (n=6) GMTs did not change appreciatively postvaccination over time.

### 4.3 Safety

The most frequently reported treatment-related systemic solicited AEs were fatigue, headache, and myalgia, each occurring in more than 15% of participants in the CHIKV VLP vaccine group (**Table 3**). The most common treatment-related local solicited AE was injection site pain, reported by 24% of participants in the vaccine group. On average, duration of treatment-related solicited AEs in the vaccine group was 2.0 days, with onset for most on Day 1 or Day 2. Treatment-related unsolicited AEs were reported by 2.3% of participants in the vaccine group and by 1.5% of participants in the placebo group, most of which were grade 1 or 2 in severity (Table S4 in the Supplementary Appendix). The single incident of grade 3 treatment-related unsolicited AE was a case of dehydration which resolved without medical intervention in the vaccine group. The most common treatment-related unsolicited AEs that occurred beyond the 7 day solicited event collection window were headache (vaccine: 0.3%, placebo: 0.2%) and arthralgia (vaccine: 0.3%, placebo: 0.4%). Notably, more females reported solicited AEs in both the vaccine and placebo groups (vaccine: 46%, placebo: 31%), compared to males (vaccine: 30%, placebo: 24%).

**Table 3.**
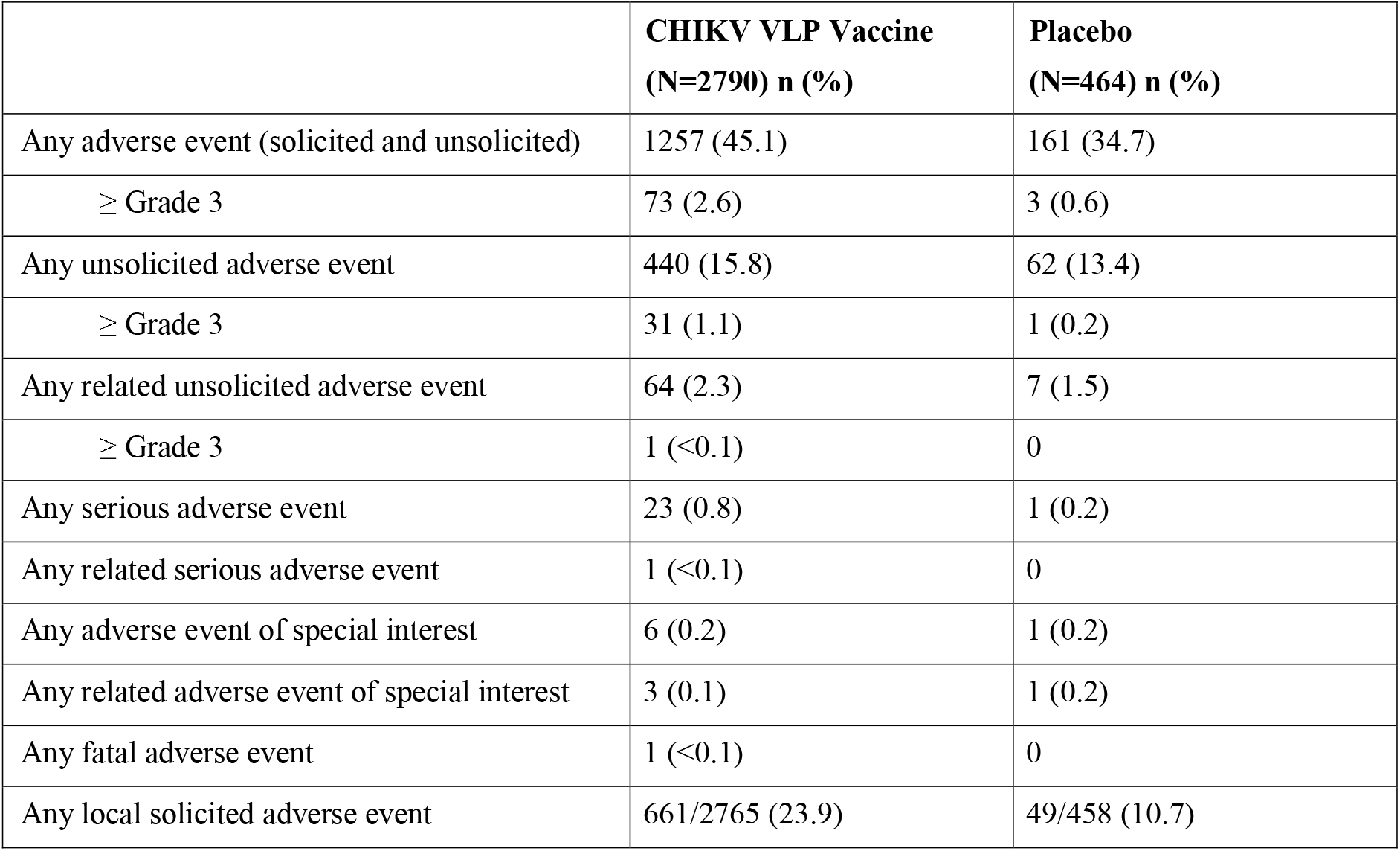
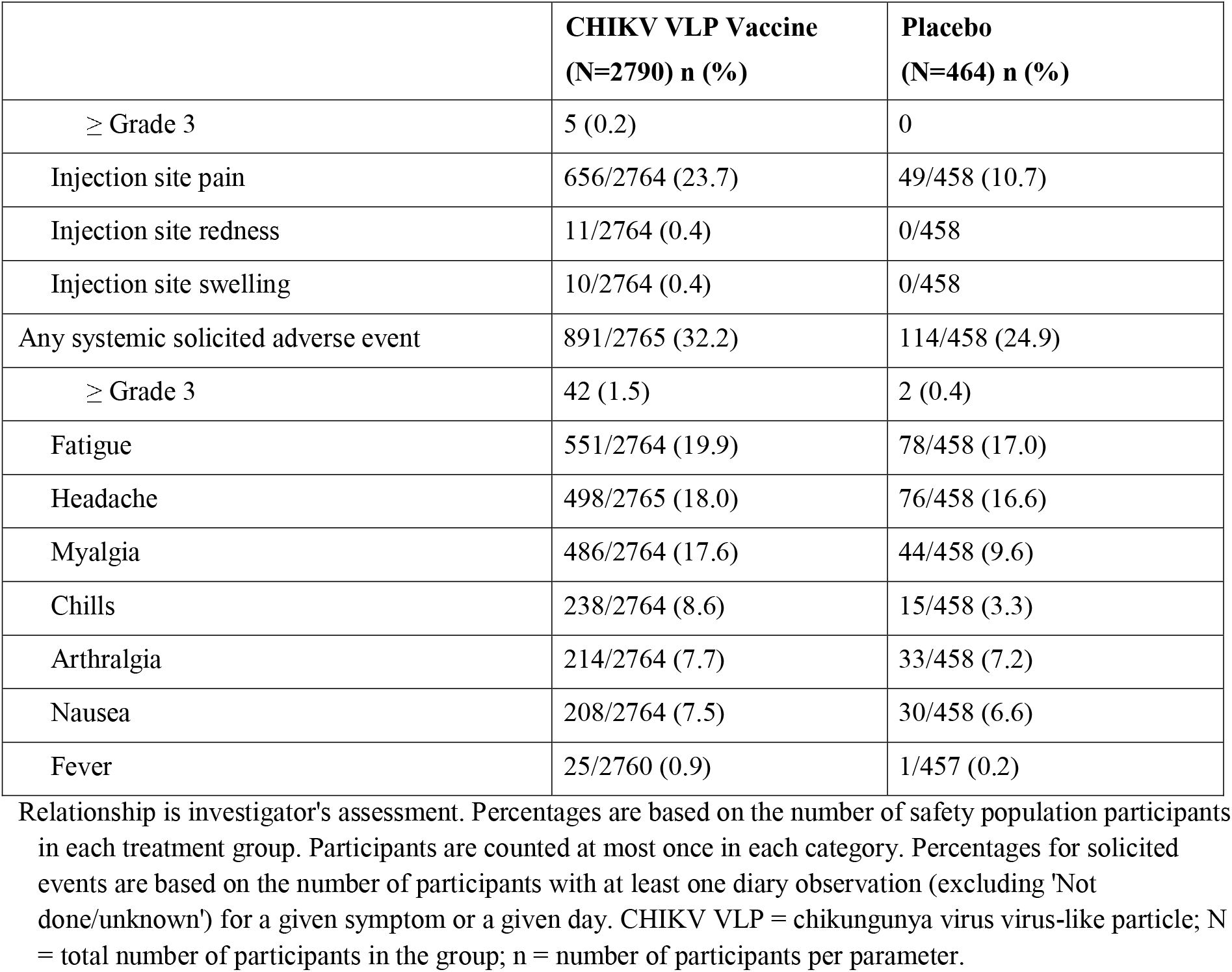
Summary of Adverse Events (Safety Population).

|  | **CHIKV VLP Vaccine**<br>**(N=2790) n (%)** | **Placebo**<br>**(N=464) n (%)** |
| --- | --- | --- |
| Any adverse event (solicited and unsolicited) | 1257 (45.1) | 161 (34.7) |
| ≥ Grade 3 | 73 (2.6) | 3 (0.6) |
| Any unsolicited adverse event | 440 (15.8) | 62 (13.4) |
| ≥ Grade 3 | 31 (1.1) | 1 (0.2) |
| Any related unsolicited adverse event | 64 (2.3) | 7 (1.5) |
| ≥ Grade 3 | 1 (<0.1) | 0 |
| Any serious adverse event | 23 (0.8) | 1 (0.2) |
| Any related serious adverse event | 1 (<0.1) | 0 |
| Any adverse event of special interest | 6 (0.2) | 1 (0.2) |
| Any related adverse event of special interest | 3 (0.1) | 1 (0.2) |
| Any fatal adverse event | 1 (<0.1) | 0 |
| Any local solicited adverse event | 661/2765 (23.9) | 49/458 (10.7) |
| ≥ Grade 3 | 5 (0.2) | 0 |
| Injection site pain | 656/2764 (23.7) | 49/458 (10.7) |
| Injection site redness | 11/2764 (0.4) | 0/458 |
| Injection site swelling | 10/2764 (0.4) | 0/458 |
| Any systemic solicited adverse event | 891/2765 (32.2) | 114/458 (24.9) |
| ≥ Grade 3 | 42 (1.5) | 2 (0.4) |
| Fatigue | 551/2764 (19.9) | 78/458 (17.0) |
| Headache | 498/2765 (18.0) | 76/458 (16.6) |
| Myalgia | 486/2764 (17.6) | 44/458 (9.6) |
| Chills | 238/2764 (8.6) | 15/458 (3.3) |
| Arthralgia | 214/2764 (7.7) | 33/458 (7.2) |
| Nausea | 208/2764 (7.5) | 30/458 (6.6) |
| Fever | 25/2760 (0.9) | 1/457 (0.2) |

Relationship is investigator's assessment. Percentages are based on the number of safety population participants in each treatment group. Participants are counted at most once in each category. Percentages for solicited events are based on the number of participants with at least one diary observation (excluding 'Not done/unknown') for a given symptom or a given day. CHIKV VLP = chikungunya virus virus-like particle; N = total number of participants in the group; n = number of participants per parameter.

SAEs were reported by 23 (0.8%) vaccine recipients and one placebo participant (0.2%). One SAE, a retinal detachment in a 56-60-year-old female, was assessed as possibly related to trial treatment by the site investigator but was assessed by the sponsor and independent Safety Monitoring Committee Chair as unrelated due to prior medical history of seeing ‘black spots’ in the affected right eye 1 month before the trial.

Adverse events of special interest, defined as new onset or worsening arthralgia that was medically attended, were reported by 7 participants, 6 in the vaccine group (0.2%) and 1 in the placebo group (0.2%); all AESI were grade 1 or 2 in severity. Four AESI were assessed as related to trial treatment (3 arthralgia in the vaccine group [0.1%] and 1 arthralgia in the placebo group [0.2%]). Treatment-related MAAEs were reported by 14 (0.5%) vaccine recipients and 2 (0.4%) placebo recipients, all of which were grade 1 or 2 in severity.

There was one death in the vaccine group unrelated to trial treatment. Two participants in the vaccine group became pregnant during the trial despite precautionary measures taken to prevent pregnancy as detailed in the inclusion criteria. One participant, a 26-30-year-old female, became pregnant 5.5 months after being vaccinated. Her prenatal care was complicated by gestational hypertension and diabetes and was managed by a high-risk Maternal Fetal Medicine Physician. The baby had fronto-ethmoid encephalocele; this fetal abnormality was assessed as unrelated to trial vaccine. The second pregnancy occurred in a 36-40-year-old female, 1 month and 27 days after vaccine administration, and resulted in the birth of a healthy infant.

## 5 DISCUSSION

In this pivotal phase 3 trial, the VLP-based chikungunya vaccine was immunogenic in healthy adolescents and adults aged 12 to 64 years, as demonstrated by 96.8% of participants reaching seroprotective levels of neutralizing antibodies at Day 15 (14 days postvaccination) and 97.8% at Day 22 (21 days postvaccination). Rapid immune response as early as 1 week post-injection was seen with nearly half of participants showing a SRR of 46.6% at Day 8. This early response, however, was more pronounced in adolescents who had Day 8 SRR of 70.9%, compared to adults where just over half of the 18 to 45-year-old group reached the seroresponse threshold (51.2%), and 33.4% of the 46 to 64-year-old group. Durability of antibody response was demonstrated up to 6-months postvaccination (overall Day 183 SRR of 85.5%). The durability was higher in the adolescent group (94.8%) compared to the two adult subgroups (18 to 45-year-old group: 85.0% and 46 to 64-year-old group: 84.1%). Another phase 3 trial (NCT05349617) confirmed the immunogenicity and durability in individuals ≥65 years of age. The trend of successively lower immune responses from younger to older age groups is expected as immune function is known to decline with age. In a systematic review of infectious disease outcomes, it was suggested that peak immune function may occur at age 5 to 14 years, followed by a rise in infectious disease severity (decreasing immunity) by age 20 for many infections, and a steeper rise of infections (further decreasing immunity) into old age, although the pattern varies by disease.^14^

Post ad hoc analysis of 63 participants who received the CHIKV VLP vaccine but had detectable amounts of CHIKV neutralizing antibodies at baseline found that baseline seropositive vaccine recipients had notably higher SNA titers compared to baseline seronegative vaccine recipients at all timepoints. Despite the small sample size of this analysis, these results provide promising evidence that CHIKV VLP vaccine will be well tolerated and elicit a strong boost response in individuals with previous CHIKV infection.

CHIKV VLP vaccine has a favorable safety profile, with most AEs being grade 1 or 2 in severity and self-limiting with resolution within 2 days. The most frequently reported treatment-related solicited AEs were fatigue, headache, myalgia, and injection site pain. SAEs were infrequent (<1%) and the incidence of AESI and MAAEs did not differ between the vaccine and placebo groups.

A limitation of this trial is that the determination of CHIKV VLP vaccine effectiveness was based on an immunological surrogate-marker. The immunogenicity threshold used in this trial was defined based on a nonhuman primate passive antibody transfer study where human sera from CHIKV VLP vaccine vaccinated individuals was used to prevent viremia following wild-type CHIKV challenge in cynomolgus macaques. In nonhuman primates, an SNA titer of 23.6 was estimated to be associated with an 80% probability of protection from CHIKV. Based on those results, an SNA titer of 100 was conservatively established as the protective threshold in humans (FDA/EMA agreed threshold). Efficacy against CHIKV disease will be investigated in a future clinical endpoint trial to be conducted in CHIKV endemic regions.

Another limitation is that the trial population included healthy individuals; further exploration may be useful to assess specific disease populations. Additionally, due to the limited number of pregnant participants, the safety of CHIKV VLP vaccine in pregnant and lactating participants could not be established. There were no apparent vaccine pregnancy-related safety signals in the 11 pregnancies across the CHIKV VLP vaccine clinical development program nor in the 2 developmental and reproductive toxicology studies performed in rabbits and rats. Finally, our immunogenicity threshold is specific to the validated CHIKV human SNA assay used in this trial and, thus, cannot be directly compared with thresholds established using other assays.

CHIKV VLP vaccine is based on the VLP platform. Virus-like particles are assembled from viral structural proteins that conformationally resemble wild-type virions but lack genetic material preventing replication in the host, which reduces the risk of unexpected adverse effects or mutations vs live-attenuated virus vaccines.^15,16,17,18^ VLP vaccines are nonreplicating, have been widely used for over 30 years, and generally are not contraindicated for use in immunocompromised populations and pregnant women.

CHIKV VLP vaccine offers the first VLP-based vaccine option for immunization against CHIKV disease, providing a potential safe alternative to participants in whom live vaccines are contraindicated. Additionally, CHIKV VLP vaccine is the first CHIKV vaccine with safety and immunogenicity data in adolescents 12 years of age and older, providing critical access to chikungunya vaccines for this age group. Findings from this pivotal phase 3 trial demonstrate that CHIKV VLP vaccine induces a rapid and robust immune response as early as 7 days and through at least 6 months postvaccination, with a favorable safety profile.

## Data Availability

De-identified data produced in the present study are available upon reasonable request to the authors.

## 6. ACKNOWLEDGEMENTS

We thank all participants, investigators, and trial site personnel who took part in this clinical trial. We also thank all employees of Bavarian Nordic A/S and Emergent BioSolutions Inc. past and present who were involved, as well as members of the Safety Monitoring Committee.

