## Supplementary Appendix for "Chikungunya Virus VLP Vaccine: Phase 3 Trial in Adolescents and Adults"

John Ervin, MD, Alliance for Multispecialty Research – Kansas City, Kansas City, MO

Carlos Fierro, MD, Johnson County ClinTrials, Lenexa, KS

Terry Klein, MD, Alliance for Multispecialty Research – Wichita East, Wichita, KS

Cayce Tangeman, MD, Coastal Carolina Research Center, North Charleston, SC

Cynthia Becher Strout, MD, Coastal Carolina Research Center, North Charleston, SC

Harry Studdard, MD, Alliance for Multispecialty Research – Mobile, Mobile, AL

Matthew Davis, MD, Rochester Clinical Research, Inc., Rochester, NY

Barbara Rizzardi, MD, Advanced Clinical Research, West Jordan, UT

Margaret Rhee, MD, Velocity Clinical Research, Inc., Cleveland, OH

Joan Rothenberg, MD, Velocity Clinical Research, Inc., Cleveland, OH

Samir Arora, MD, Aventiv Research Inc., Columbus, OH

Mahashweta Rita Ghosh, MD, Optimal Research, LLC, Rockville, MD

Wayne Farnsworth, MD, Optimal Research, LLC, Rockville, MD

Peta Gay Jackson-Booth, MD, Optimal Research, LLC, Rockville, MD

Corey G. Anderson, MD, Alliance for Multispecialty Research, LLC, Tempe, AZ

Steven Cox, DO, Lynn Institute of Norman, Norman, OK

Michael L. Levin, MD, WR-CRCN, LLC, Las Vegas, NV

Liliana Ruiz-Leon, DO, WR-CRCN, LLC, Las Vegas, NV

Marian E. Shaw, MD, Velocity Clinical Research, Boise, Meridian, ID

Mark Alan Turner, MD, Velocity Clinical Research, Boise, Meridian, ID

Mira Baron, MD, Palm Beach Research Center, West Palm Beach, FL

Robert Frenck, Jr., MD, Cincinnati Children’s Hospital Medical Center – The Gamble Vaccine Research Center, Cincinnati, OH

David L. Fried, MD, Velocity Clinical Research – Providence, East Greenwich, RI

Carl Griffin, MD, Lynn Health Science Institute, Oklahoma City, OK

Ripley Hollister, MD, Lynn Institute of the Rockies, Colorado Springs, CO

Muhammad Irfan, MD, DM Clinical Research, Tomball, TX

Michael Jacobs, MD, Alliance for Multispecialty Research, LLC, Las Vegas, NV

Jeffry A. Jacqmein, MD, Jacksonville Center for Clinical Research, Jacksonville, FL

Vicki Miller, MD, Texas Center for Drug Development, Inc., Houston, TX

Mark S. Adams, MD, Alliance for Multispecialty Research, LLC, Lexington, KY

Sarah George, MD, Saint Louis University, Saint Louis, MO

Bruce G. Rankin, DO, Accel Research Sites – DeLand Clinical Research Unit, Lake Mary, FL

Judith L. Kirstein, MD, Velocity Clinical Research, Banning, Banning, CA

Mary Bailey, MD, Alliance for Multispecialty Research, LLC, Norfolk, VA

Richard M. Glover, II, MD, Alliance for Multispecialty Research, LLC, Newton, KS

Sinikka Liisa Green, Velocity Clinical Research, Austin, Cedar Park, TX

Gregg Lucksinger, MD, Velocity Clinical Research, Austin, Cedar Park, TX

Lisa M. Cohen, DO, M3 Wake Research, Inc., Raleigh, NC

James Kopp, MD, Synexus Clinical Research US, Inc., Anderson, SC

Linda Murray, DO, Synexus Clinical Research US, Inc., Pinellas Park, FL

Joseph Newberg, MD, Synexus Clinical Research US, Inc., Chicago, IL

Ramon G. Reyes (Almodovar), MD, BFHC Research, San Antonio, TX

William B. Smith, MD, Alliance for Multispecialty Research, LLC, Knoxville, TN

Leslie M. Tharenos, MD, Synexus Clinical Research US, Inc., St. Louis, MO

Robert Noveck, MD, Alliance for Multispecialty Research, LLC, New Orleans, LA

Ramon Vargas, MD, Alliance for Multispecialty Research, LLC, New Orleans, LA

William Jones, MD, Trial Management Associates, LLC, Wilmington, NC

Daniel H. Brune, MD, Optimal Research LLC, Peoria, IL

Murray A. Kimmel, DO, Optimal Research, LLC, Melbourne, FL

Randle T. Middleton, MD, Optimal Research, LLC, Huntsville, AL

Patrick R. Yassini, MD, Optimal Research, LLC, San Diego, CA

Sarah Smiley, DO, Velocity Clinical Research, Medford, Medford, OR

Jeffrey Adelglass, MD, Research Your Health, Plano, TX

Jorge T. Caso, MD, Suncoast Research Associates, LLC, Miami, FL

Safety Monitoring Committee

Chairs

Julie Ledgerwood, DO (until Jul-2022)

Vaccine Research Center and National Institute of Allergy and Infectious Diseases, National Institutes of Health

David R. Snydman, MD, FACP, FIDSA, FAST (Nov-2022 onwards)

Tufts Medical Center and Tufts University School of Medicine

Members

Michelle Petri, MD MPH

Johns Hopkins University School of Medicine and Johns Hopkins Hospital

Ying Lu, PhD

Stanford University School of Medicine

Supplementary methods

Inclusion and exclusion criteria

Inclusion Criteria

Participants were required to meet **all** the following criteria to be enrolled:

1. Able and willing to provide informed consent (and assent, as applicable) voluntarily signed by participant (and guardian, as applicable).
2. Males or females, 12 to <65 years of age.
3. Generally healthy, in the opinion of the investigator, based on medical history, physical examination, and screening laboratory assessments.
4. Women who were either:
a. Not of childbearing potential (CBP): premenarche, surgically sterile (at least six weeks post bilateral tubal ligation, bilateral oophorectomy, or hysterectomy), or post-menopausal (defined as a history of ≥12 consecutive months without menses prior to randomization in the absence of other pathologic or physiologic causes, following cessation of exogenous sex-hormonal treatment).

**or**:

b. Meeting all the below criteria:

- Negative serum pregnancy test at Screening Visit.
- Negative urine pregnancy test immediately prior to dosing at Day 1.
- Use one of these acceptable methods of contraception (if women of CBP) for the duration of participation:
- Hormonal contraceptives (eg, implants, pills, patches) initiated ≥30 days prior to dosing.
- Intrauterine device (IUD) inserted ≥30 days prior to dosing.
- Double barrier type of birth control (male condom with female diaphragm, male condom with cervical cap).
- Abstinence was acceptable only for adolescents (12 to <18 years old) who were not sexually active.

Note: See protocol (online at ClinicalTrials.gov, NCT05072080) for this trial's list of acceptable methods of contraception.

Note: Contraception requirements did not apply for participants in exclusively same-sex relationships and these participants should have had no plans to become pregnant by any other means for the duration of the trial.

Exclusion Criteria

Participants who met **any** of the following criteria **could not** be enrolled:

1. Currently pregnant, breastfeeding, or planning to become pregnant during the trial.
2. Body mass index (BMI) ≥35 kg/m^2^.
3. Positive laboratory evidence of current infection with Human Immunodeficiency Virus HIV-1/HIV-2, Hepatitis C Virus, or Hepatitis B Virus.
4. History of severe allergic reaction or anaphylaxis to any component of the IP.
5. History of any known congenital or acquired immunodeficiency that could impact response to vaccination (eg, leukemia, lymphoma, generalized malignancy, functional or anatomic asplenia, alcoholic cirrhosis).
6. Prior receipt or anticipated use of systemic immunomodulatory or immunosuppressive medications from six months prior to screening through Day 22. Note: For systemic corticosteroid use at a dose or equivalent dose of 20 mg of prednisone daily for 14 days or more within three months of screening through Day 22 was exclusionary. The use of inhaled, intranasal, topical, ocular, or intraocular steroids was allowed.
7. Receipt or anticipated receipt of blood or blood-derived products from 90 days prior to screening through Day 22.
8. Acute disease within the last 14 days (participants with an acute mild febrile illness could be considered for a deferral of vaccination two weeks after the illness had resolved and treatment had been completed).
9. Clinically significant cardiac, pulmonary, rheumatologic, or other chronic disease, in the opinion of the investigator. This may include chronic illness requiring hospitalization in the last 30 days prior to screening.
10. Enrollment in an interventional study and/or receipt of another investigational product from 30 days prior to screening through the duration of study participation.
11. Receipt or anticipated receipt of any vaccine from 30 days prior to Day 1 through Day 22.
12. Evidence of substance abuse that, in the opinion of the investigator, could adversely impact the participant's participation or the conduct of the trial.
13. Prior receipt of an investigational CHIKV vaccine/product.
14. Any other medical condition that, in the opinion of the investigator, could adversely impact the participant's participation or the conduct of the trial.

### CHIKV human serum neutralizing antibody (SNA) assay

A Chikungunya virus neutralization assay (CHIKV human SNA assay) was used to measure serum neutralizing antibody titers to CHIKV in trial participants. The CHIKV human SNA assay is based on capacity of serum antibodies to neutralize recombinant CHIKV 181/25 virus (attenuated derivative of Asian strain AF15561) engineered to express luciferase (CHIKV-*luc*). Reductions of luciferase activity are quantified in cultures of Vero cells exposed to mixtures of CHIKV-*luc* virus and dilutions of test serum. Quantitation of reporter gene expression correlating to the level of CHIKV-*luc* virus infected cells, is determined by detection of luciferase activity in assay wells using luciferin substrate and a microplate luminometer to measure luminescence. The CHIKV antibody 80% neutralizing titer (NT_80_), calculated using linear regression and interpolation analysis, is the reciprocal of the maximum serum dilution that provides 80% protection of Vero cells from CHIKV-*luc* infection (80% reduction of luciferase activity compared to virus only control).

Supplementary statistical analysis

Coprimary endpoint: Day 22 seroresponse rate

The superiority of the immune response to CHIKV VLP vaccine over that to placebo was demonstrated at Day 22 by comparing seroresponse rates between the two treatment groups (the proportion of participants with a human SNA assay ≥100; considered the presumptive seroprotection rate). The difference in seroresponse rates between CHIKV VLP vaccine and placebo groups was calculated, along with the 95% CI for the difference based on the Newcombe hybrid score method. The lower bound of the two-sided 95% CI on the difference in seroresponse rates between CHIKV VLP vaccine and placebo groups had to be ≥70%. Additionally, the null hypothesis of no difference between seroresponse rate proportions was tested using a chi-square test with alpha=0.05. No multiplicity adjustment was employed and no covariate adjustment was performed.

The primary comparison was in the IEP across all age groups combined. Tests were repeated in the modified intent-to-treat (mITT) population as a measure of robustness, along with tests for each population in the separate age strata. No multiplicity adjustment was made for the analysis of the separate age strata as the primary population was the combined age groups.

Coprimary endpoint: Day 22 geometric mean titer

Day 22 GMTs were compared between CHIKV VLP vaccine and placebo treatment groups and were analyzed via a linear model based on an alpha=0.05. The primary model was an ANOVA, with logarithmically transformed CHIKV SNA titers (log10) as the dependent variable and treatment group and trial site as the fixed effects in the model. The adjusted least square means and their 95% CIs calculated based on the ANOVA were back transformed and reported as the group GMT values. All tests were carried out at a two-sided significance level of 0.05 and no adjustment for multiplicity was applied.

The primary comparison was in the IEP across all age groups combined. Tests were repeated in the mITT population as a measure of robustness, along with tests for each population in the separate age strata. No multiplicity adjustment was made for the analysis of the separate age strata as the primary population was the combined age groups.

Coprimary endpoint: Day 22 lot consistency

The consistency of the immune response across three consecutively manufactured lots of CHIKV VLP vaccine was confirmed at Day 22. Immunogenicity as measured by human SNA assay CHIKV SNA GMTs were compared using ratios of GMTs between all three pairs of CHIKV VLP vaccine lots (A:B, B:C, A:C) for the IEP in the 18 to 45 years of age stratum, and the analysis was repeated for the mITT population as a measure of the robustness of the results.

The GMTs associated with the primary lot consistency objective were calculated via a linear model for the CHIKV SNA titers collected at Day 22. The primary model was an ANOVA, with log10-transformed CHIKV SNA titers as the dependent variable and vaccine lot and trial site as the fixed effects in the model. The adjusted least square means and their 95% CIs calculated based on the ANOVA was back transformed and reported as the vaccine lot GMT values. The ratio of GMTs for the comparison of the three pairs of CHIKV VLP vaccine lots (A:B, B:C, A:C) was derived from this model.

All tests were carried out at a two-sided significance level of 0.05 and no adjustment for multiplicity was applied. If for all pairs of vaccine lots, the two-sided 95% CI of the GMT ratio was within [0.667; 1.5] then lot-to-lot consistency was demonstrated.

Key secondary endpoint: Seroresponse rate at Days 15, 183, and 8, in that order

Seroresponse rates and seroresponse rate differences (CHIKV VLP vaccine minus placebo) with associated 95% CIs based on antibody titers measured at Days 15, 183, and 8 was analyzed as described above for Day 22. For each timepoint, the null hypothesis of no difference between seroresponse rate proportions in the CHIKV VLP vaccine vs placebo group was tested using a chi square test with alpha=0.05. For Day 15 only, the lower bound of the two-sided 95% CI on the difference in seroresponse rates between CHIKV VLP vaccine and placebo was compared to ≥70% for assessment of clinical significance.

A prespecified hierarchical approach was employed for the key secondary immunogenicity endpoint hypothesis testing to preserve the type I error rate without the need for further multiplicity adjustment. If the null hypotheses were rejected for all coprimary endpoints, only then were the key secondary endpoints formally tested in sequential order. If a nonsignificant test was reached formal testing was to stop, and the remaining endpoints would be reported for information only. The seroresponse rates and rate differences at Days 15, 183, and 8 were tested sequentially in order, each for the IEP across both age groups combined.

After testing the coprimary immunogenicity endpoints and, if all coprimary were met the hierarchical key secondary immunogenicity endpoints, no other formal hypothesis testing was carried out. The remaining secondary immunogenicity endpoints including GMTs, GMFI, other titers, the exploratory endpoint, and all safety endpoints were evaluated and reported for information only, thus no further multiplicity adjustment was needed.

Secondary endpoint: Geometric mean titer at Days 15, 183, and 8

For the comparison of CHIKV VLP vaccine to placebo, GMTs based on antibody titers measured at Days 8, 15, and 183 were analyzed as described above for Day 22. Geometric mean fold increases for increase over Day 1 titer was analyzed as described for GMTs for each postvaccination timepoint.

Secondary endpoint: Seroconversion at other titers

As described above, secondary response rates at other titers (eg, 15 and 4-fold rise over baseline) were reported with associated two-sided 95% Wilson method CIs by scheduled visit for each treatment group.

Supplementary figures

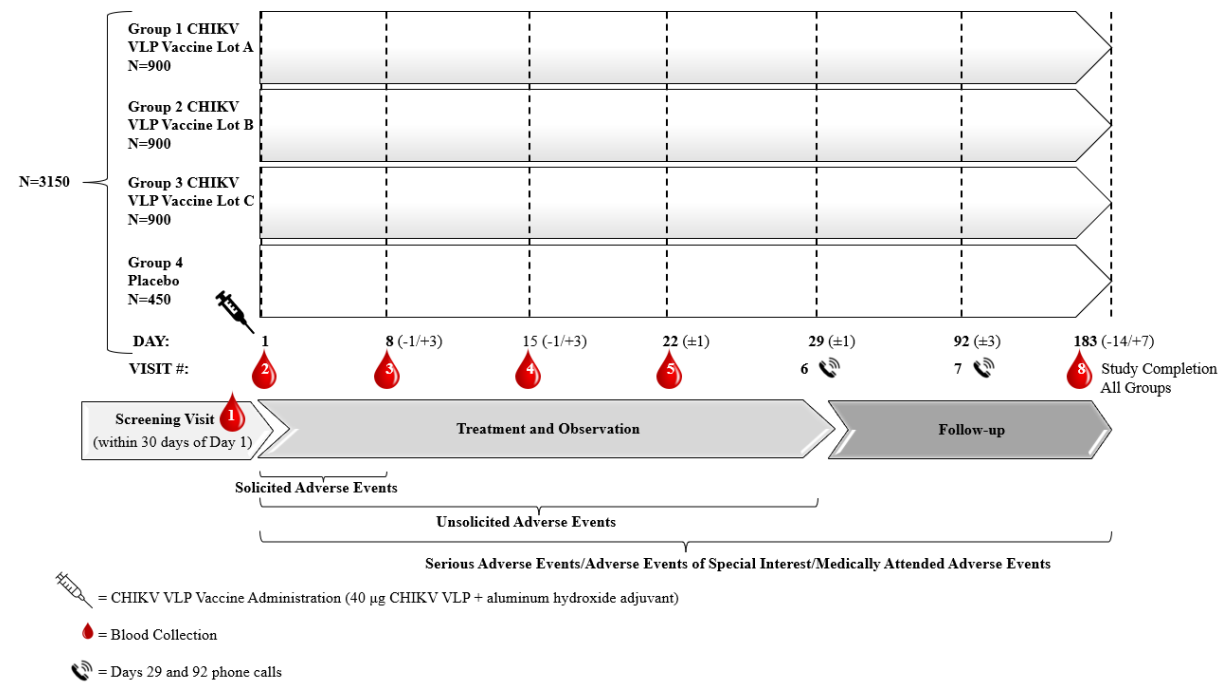

Figure S 1	Trial Design.

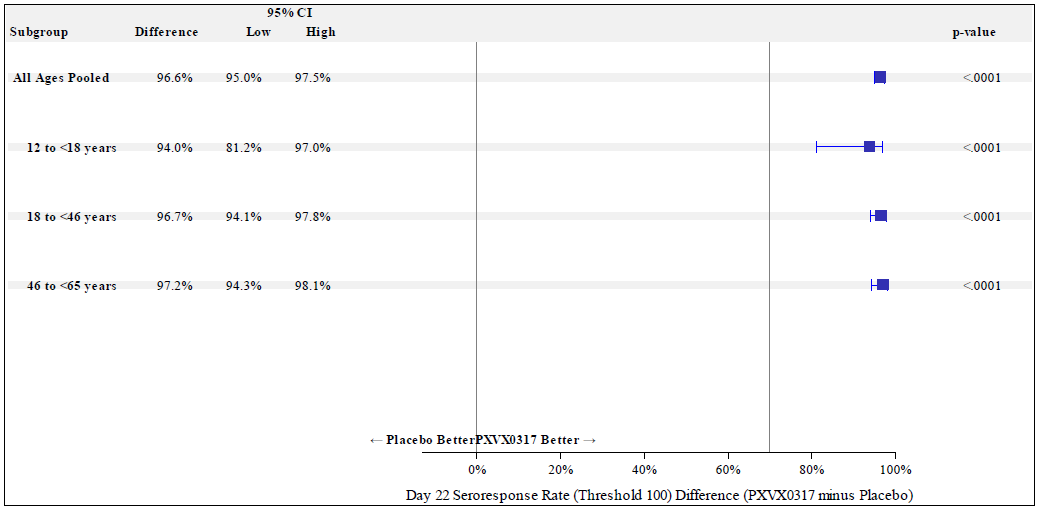

Note: Error bars are 95% CI based on the Newcombe hybrid score method; p-values are from a two-sided chi-square test of equality of seroresponse percentages between groups. CI = confidence interval; PXVX0317 = CHIKV VLP vaccine.

Figure S 2	Forest Plot of Seroresponse Rate Differences at Day 22 (Immunogenicity Evaluable Population, All Ages Pooled and by Age Group).

Supplementary tables

Table S 1	CHIKV SNA geometric mean titer and geometric mean fold increase from baseline by visit (immunogenicity evaluable population)

| Day  Statistic | CHIKV VLP Vaccine  (N=2559) | Placebo  (N=424) | Ratio of GMTs  [95% CI]^a^ | P value^c,e^ |
| --- | --- | --- | --- | --- |
| **Day 1** | | | | |
| n^b^ | 2559 | 424 |  |  |
| GMT^c^ [95% CI]  Median^d^  Min, Max^d^ | 7.50 [7.50, 7.50]  7.50  7.5, 7.5 | 7.50 [7.50, 7.50]  7.50  7.5, 7.5 | - | - |
| **Day 8** | | | | |
| n^b^ | 2510 | 419 |  |  |
| GMT^c^ [95% CI]  Median^d^  Min, Max^d^ | 93.36 [87.17, 99.99]  90.55  7.5, 48641.3 | 7.40 [6.49, 8.44]  7.50  7.5, 9949.3 | 12.62 [11.06, 14.41] | <0.0001 |
| GMFI^e^ [95% CI]  Median^d^  Min, Max^d^ | 12.45 [11.62, 13.33]  6.04  1.0, 3242.8 | 0.99 [0.86, 1.13]  1.00  1.0, 663.3 | - | <0.0001 |
| **Day 15** | | | | |
| n^b^ | 2434 | 395 |  |  |
| GMT^c^ [95% CI]  Median^d^  Min, Max^d^ | 1095.83 [1029.28, 1166.69]  1203.90  7.5, 52909.3 | 7.62 [6.76, 8.59]  7.50  7.5, 1267.5 | 143.88 [127.60, 162.24] | <0.0001 |
| GMFI^e^ [95% CI]  Median^d^  Min, Max^d^ | 146.11 [137.24, 155.56]  80.26  1.0, 3527.3 | 1.02 [0.90, 1.14]  1.00  1.0, 84.5 | - | <0.0001 |
| **Day 22** | | | | |
| n^b^ | 2559 | 424 |  |  |
| GMT^c^ [95% CI]  Median^d^  Min, Max^d^ | 1618.05 [1522.11, 1720.04]  1787.30  7.5, 60115.8 | 7.85 [6.98, 8.83]  7.50  7.5, 6007.3 | 206.12 [183.17, 231.95] | <0.0001 |
| GMFI^e^ [95% CI]  Median^d^  Min, Max^d^ | 215.74 [202.95, 229.34]  119.15  1.0, 4007.7 | 1.05 [0.93, 1.18]  1.00  1.0, 400.5 | - | <0.0001 |
| **Day 183** | | | | |
| n^b^ | 2301 | 401 |  |  |
| GMT^c^ [95% CI]  Median^d^  Min, Max^d^ | 337.73 [318.27, 358.37]  379.50  7.5, 9205.6 | 8.19 [7.33, 9.14]  7.50  7.5, 583.5 | 41.26 [36.95, 46.07] | <0.0001 |
| GMFI^e^ [95% CI]  Median^d^  Min, Max^d^ | 45.03 [42.44, 47.78]  25.30  1.0, 613.7 | 1.09 [0.98, 1.22]  1.00  1.0, 38.9 | - | <0.0001 |
| CI = confidence interval; GMT = geometric mean titer; GMFI = geometric mean fold increase; LLOQ = lower limit of quantitation; Max = maximum; Min = minimum; SNA = serum neutralizing antibody.  Note: Values below lower limit of quantitation (LLOQ=15) were assigned the value LLOQ/2=7.5.  ^a^ Ratio of GMTs is (vaccine:placebo).  ^b^ n is the number of participants with a sample result available at the indicated visit.  ^c^ Geometric mean titer estimates, together with their 95% CIs, are derived from an ANOVA model that includes site and treatment group as fixed effects, assuming normality of the log titers. Ratio of GMTs and 95% CIs are derived from the same model. p-value tests equivalence of group GMTs on the log scale (ie, ratio of GMTs equal to 1).  ^d^ Median and range are based on raw values with imputation per note.  ^e^ Geometric mean fold increase from Day 1 (baseline) is (Day x/Day 1). GMFI estimates and 95% CIs are based on t-statistics assuming a normal distribution of the log fold increase in titer. p-value tests equality of log fold increase in titer between groups. | | | | |

Table S 2	CHIKV SNA seroresponse rate by visit, sex, race, ethnicity, and age (immunogenicity evaluable population)

| Day | Group | Subgroup | Seroresponse CHIKV VLP Vaccine (N=2559)  **n/N (%)^c^ [95% CI]^d^** | Seroresponse Placebo (N=424)  **n/N (%)^c^ [95% CI]^d^** | Seroresponse Rate Difference [95% CI]^a,b^ | P value |
| --- | --- | --- | --- | --- | --- | --- |
| **Day 8** | All IEP | All IEP | 1169/2510 (46.6) [44.6, 48.5] | 2/419 (0.5) [0.1, 1.7] | 46.1 [43.8, 48.1] | <0.0001 |
|  | Sex | Male | 516/1216 (42.4) [39.7, 45.2] | 1/207 (0.5) [0.1, 2.7] | 42.0 [38.4, 44.8] | <0.0001 |
|  |  | Female | 653/1294 (50.5) [47.7, 53.2] | 1/212 (0.5) [0.1, 2.6] | 50.0 [46.5, 52.7] | <0.0001 |
|  | Race | White | 862/1853 (46.5) [44.3, 48.8] | 2/313 (0.6) [0.2, 2.3] | 45.9 [43.1, 48.2] | <0.0001 |
|  |  | Non-White | 297/638 (46.6) [42.7, 50.4] | 0/102 (0.0) [0.0, 3.6] | 46.6 [41.3, 50.4] | <0.0001 |
|  | Ethnicity | Non-Hispanic or Latino | 927/2013 (46.1) [43.9, 48.2] | 1/343 (0.3) [ 0.1, 1.6] | 45.8 [ 43.2, 48.0] | <0.0001 |
|  |  | Hispanic or Latino | 217/444 (48.9) [44.3, 53.5] | 1/64 (1.6) [ 0.3, 8.3] | 47.3 [ 39.1, 52.1] | <0.0001 |
|  | Age | 12 to <18 years | 141/199 (70.9) [64.2, 76.7] | 1/33 (3.0) [0.5, 15.3] | 67.8 [53.8, 74.2] | <0.0001 |
|  |  | 18 to <46 years | 737/1440 (51.2) [48.6, 53.8] | 1/242 (0.4) [0.1, 2.3] | 50.8 [47.6, 53.4] | <0.0001 |
|  |  | 46 to <65 years | 291/871 (33.4) [30.4, 36.6] | 0/144 (0.0) [0.0, 2.6] | 33.4 [29.4, 36.6] | <0.0001 |
| **Day 15** | All IEP | All IEP | 2355/2434 (96.8) [96.0, 97.4] | 3/395 (0.8) [0.3, 2.2] | 96.0 [94.3, 96.8] | <0.0001 |
|  | Sex | Male | 1132/1182 (95.8) [94.5, 96.8] | 2/197 (1.0) [0.3, 3.6] | 94.8 [91.8, 96.0] | <0.0001 |
|  |  | Female | 1223/1252 (97.7) [96.7, 98.4] | 1/198 (0.5) [0.1, 2.8] | 97.2 [94.7, 98.0] | <0.0001 |
|  | Race | White | 1736/1799 (96.5) [95.5, 97.3] | 3/291 (1.0) [0.4, 3.0] | 95.5 [93.3, 96.5] | <0.0001 |
|  |  | Non-White | 599/615 (97.4) [95.8, 98.4] | 0/100 (0.0) [0.0, 3.7] | 97.4 [93.4, 98.4] | <0.0001 |
|  | Ethnicity | Non-Hispanic or Latino | 1877/1943 (96.6) [95.7, 97.3] | 1/323 (0.3) [ 0.1, 1.7] | 96.3 [ 94.6, 97.1] | <0.0001 |
|  |  | Hispanic or Latino | 427/440 (97.0) [95.0, 98.3] | 2/60 (3.3) [ 0.9, 11.4] | 93.7 [ 85.4, 96.4] | <0.0001 |
|  | Age | 12 to <18 years | 187/191 (97.9) [94.7, 99.2] | 1/30 (3.3) [0.6, 16.7] | 94.6 [80.9, 97.6] | <0.0001 |
|  |  | 18 to <46 years | 1375/1406 (97.8) [96.9, 98.4] | 2/228 (0.9) [0.2, 3.1] | 96.9 [94.5, 97.8] | <0.0001 |
|  |  | 46 to <65 years | 793/837 (94.7) [93.0, 96.1] | 0/137 (0.0) [0.0, 2.7] | 94.7 [91.5, 96.1] | <0.0001 |
| **Day 22** | All IEP | All IEP | 2503/2559 (97.8) [97.2, 98.3] | 5/424 (1.2) [0.5, 2.7] | 96.6 [95.0, 97.5] | <0.0001 |
|  | Sex | Male | 1208/1241 (97.3) [96.3, 98.1] | 3/210 (1.4) [0.5, 4.1] | 95.9 [93.0, 97.1] | <0.0001 |
|  |  | Female | 1295/1318 (98.3) [97.4, 98.8] | 2/214 (0.9) [0.3, 3.3] | 97.3 [94.8, 98.2] | <0.0001 |
|  | Race | White | 1847/1887 (97.9) [97.1, 98.4] | 2/315 (0.6) [0.2, 2.3] | 97.2 [95.4, 98.0] | <0.0001 |
|  |  | Non-White | 636/652 (97.5) [96.1, 98.5] | 3/105 (2.9) [1.0, 8.1] | 94.7 [89.3, 96.8] | <0.0001 |
|  | Ethnicity | Non-Hispanic or Latino | 2004/2049 (97.8) [97.1, 98.4] | 5/347 (1.4) [ 0.6, 3.3] | 96.4 [ 94.3, 97.4] | <0.0001 |
|  |  | Hispanic or Latino | 447/457 (97.8) [96.0, 98.8] | 0/65 (0.0) [ 0.0, 5.6] | 97.8 [ 92.0, 98.8] | <0.0001 |
|  | Age | 12 to 17 years | 195/201 (97.0) [93.6, 98.6] | 1/33 (3.0) [0.5, 15.3] | 94.0 [81.2, 97.0] | <0.0001 |
|  |  | 18 to 45 years | 1455/1480 (98.3) [97.5, 98.9] | 4/245 (1.6) [0.6, 4.1] | 96.7 [94.1, 97.8] | <0.0001 |
|  |  | 46 to <65 years | 853/878 (97.2) [95.8, 98.1] | 0/146 (0.0) [0.0, 2.6] | 97.2 [94.3, 98.1] | <0.0001 |
| **Day 183** | All IEP | All IEP | 1967/2301 (85.5) [84.0, 86.9] | 6/401 (1.5) [0.7, 3.2] | 84.0 [81.7, 85.6] | <0.0001 |
|  | Sex | Male | 853/1097 (77.8) [75.2, 80.1] | 2/196 (1.0) [0.3, 3.6] | 76.7 [73.1, 79.2] | <0.0001 |
|  |  | Female | 1114/1204 (92.5) [90.9, 93.9] | 4/205 (2.0) [0.8, 4.9] | 90.6 [87.2, 92.4] | <0.0001 |
|  | Race | White | 1528/1727 (88.5) [86.9, 89.9] | 3/300 (1.0) [0.3, 2.9] | 87.5 [85.0, 89.0] | <0.0001 |
|  |  | Non-White | 423/556 (76.1) [72.4, 79.4] | 3/97 (3.1) [1.1, 8.7] | 73.0 [66.3, 76.9] | <0.0001 |
|  | Ethnicity | Non-Hispanic or Latino | 1583/1857 (85.2) [83.6, 86.8] | 5/329 (1.5) [ 0.7, 3.5] | 83.7 [ 81.1, 85.5] | <0.0001 |
|  |  | Hispanic or Latino | 339/392 (86.5) [82.7, 89.5] | 1/60 (1.7) [ 0.3, 8.9] | 84.8 [ 76.7, 88.1] | <0.0001 |
|  | Age | 12 to 17 years | 182/192 (94.8) [90.7, 97.1] | 0/32 (0.0) [0.0, 10.7] | 94.8 [83.3, 97.1] | <0.0001 |
|  |  | 18 to 45 years | 1098/1292 (85.0) [82.9, 86.8] | 4/229 (1.7) [0.7, 4.4] | 83.2 [79.9, 85.4] | <0.0001 |
|  |  | 46 to <65 years | 687/817 (84.1) [81.4, 86.4] | 2/140 (1.4) [0.4, 5.1] | 82.7 [78.2, 85.2] | <0.0001 |
| CI = confidence interval; IEP = immunogenicity evaluable population; SNA = serum neutralizing antibody  ^a^ Seroresponse rate difference is (vaccine minus placebo); 95% CIs are based on the Newcombe hybrid score method.  ^b^ P value is from a two-sided chi-square test of equality of seroresponse percentages between groups.  ^c^ n is the number of participants with seroresponse ≥ titer 100, divided by N, the total number of participants in the group.  ^d^ 95% CIs of seroresponse rates are based on the Wilson method. | | | | | |  |

Table S 3	Participants with CHIKV SNA titer at or above selected titers (≥15 and 4-fold rise over baseline) by treatment group and visit (immunogenicity evaluable population)

| Day  Statistic | CHIKV VLP Vaccine (N=2559) n (%) | Placebo  (N=424)  n (%) | Seroresponse Rate Difference  ^[95%^ CI]^a^ | P value^b^ |
| --- | --- | --- | --- | --- |
| **Day 8** | | | | |
| n^c^ | 2510 | 419 |  |  |
| n (%) titer ≥15  [95% CI]^d^ | 2306 (91.9)  [90.7, 92.9] | 5 (1.2)  [0.5, 2.8] | 90.7 [88.7, 91.9] | <0.0001 |
| n (%) titer ≥ 4-fold rise  [95% CI]^d^ | 1644 (65.5) [63.6, 67.3] | 3 (0.7)  [0.2, 2.1] | 64.8 [62.5, 66.7] | <0.0001 |
| **Day 15** | | | | |
| n^c^ | 2434 | 395 |  |  |
| n (%) titer ≥15  [95% CI]^d^ | 2421 (99.5)  [99.1, 99.7] | 3 (0.8)  [0.3, 2.2] | 98.7 [97.2, 99.3] | <0.0001 |
| n (%) titer ≥ 4-fold Rise  [95% CI]^d^ | 2399 (98.6) [98.0, 99.0] | 3 (0.8)  [0.3, 2.2] | 97.8 [96.3, 98.4] | <0.0001 |
| **Day 22** | | | | |
| n^c^ | 2559 | 424 |  |  |
| n (%) titer ≥15  [95% CI]^d^ | 2539 (99.2)  [98.8, 99.5] | 7 (1.7)  [0.8, 3.4] | 97.6 [95.8, 98.5] | <0.0001 |
| n (%) titer ≥ 4-fold Rise  [95% CI]^d^ | 2524 (98.6) [98.1, 99.0] | 6 (1.4)  [0.7, 3.1] | 97.2 [95.5, 98.1] | <0.0001 |
| **Day 183** | | | | |
| n^c^ | 2301 | 401 |  |  |
| n (%) titer ≥15  [95% CI]^d^ | 2279 (99.0)  [98.6, 99.4] | 9 (2.2)  [1.2, 4.2] | 96.8 [94.8, 97.9] | <0.0001 |
| n (%) titer ≥ 4-fold Rise  [95% CI]^d^ | 2138 (92.9) [91.8, 93.9] | 6 (1.5)  [0.7, 3.2] | 91.4 [89.4, 92.7] | <0.0001 |
| CI = confidence interval; IEP = Immunogenicity Evaluable Population; SNA = serum neutralizing antibody.  Note: By definition, the IEP has no measurable CHIKV SNA at Day 1, therefore Day 1 is omitted from this table.  ^a^ Seroresponse rate difference is (vaccine minus placebo); 95% CIs are based on the Newcombe hybrid score method.  ^b^ P value is from a two-sided chi-square test of equality of seroresponse percentages between groups.  ^c^ n is the number of participants with a sample result available at the indicated visit.  ^d^ 95% CIs of seroresponse rates are based on the Wilson method. | | | | |

Table S 4	Treatment-related unsolicited adverse events by preferred term and highest reported severity (safety population)

| **Preferred Term** | CHIKV VLP Vaccine* (N=2790) | | | | Placebo* (N=464) | | | |
| --- | --- | --- | --- | --- | --- | --- | --- | --- |
|  | Total  m, n (%) | Grade 1  m, n (%) | Grade 2  m, n (%) | Grade 3  m, n (%) | Total  m, n (%) | Grade 1  m, n (%) | Grade 2  m, n (%) | Grade 3  m, n (%) |
| Any Treatment-related Unsolicited AE | 84, 64 (2.3) | 60, 44 (1.6) | 23, 19 (0.7) | 1, 1 (<0.1) | 11, 7 (1.5) | 6, 3 (0.6) | 5, 4 (0.9) | 0, 0 |
| Headache | 10, 9 (0.3) | 7, 7 (0.3) | 3, 2 (<0.1) | 0, 0 | 1, 1 (0.2) | 1, 1 (0.2) | 0, 0 | 0, 0 |
| Arthralgia | 7, 7 (0.3) | 5, 5 (0.2) | 2, 2 (<0.1) | 0, 0 | 2, 2 (0.4) | 1, 1 (0.2) | 1, 1 (0.2) | 0, 0 |
| Rash | 6, 5 (0.2) | 6, 5 (0.2) | 0, 0 | 0, 0 | 0, 0 | 0, 0 | 0, 0 | 0, 0 |
| Fatigue | 4, 4 (0.1) | 4, 4 (0.1) | 0, 0 | 0, 0 | 1, 1 (0.2) | 0, 0 | 1, 1 (0.2) | 0, 0 |
| Injection site pain | 4, 4 (0.1) | 4, 4 (0.1) | 0, 0 | 0, 0 | 0, 0 | 0, 0 | 0, 0 | 0, 0 |
| Dizziness | 3, 3 (0.1) | 3, 3 (0.1) | 0, 0 | 0, 0 | 1, 1 (0.2) | 1, 1 (0.2) | 0, 0 | 0, 0 |
| Myalgia | 3, 3 (0.1) | 2, 2 (<0.1) | 1, 1 (<0.1) | 0, 0 | 1, 1 (0.2) | 1, 1 (0.2) | 0, 0 | 0, 0 |
| Nasal congestion | 3, 3 (0.1) | 2, 2 (<0.1) | 1, 1 (<0.1) | 0, 0 | 0, 0 | 0, 0 | 0, 0 | 0, 0 |
| Oropharyngeal pain | 3, 3 (0.1) | 2, 2 (<0.1) | 1, 1 (<0.1) | 0, 0 | 0, 0 | 0, 0 | 0, 0 | 0, 0 |
| *There were no grade 4 treatment-related unsolicited AEs.  AE = adverse event; Grade 1 = mild; Grade 2 = moderate; Grade 3 = severe; m=Number of events, n=Number of participants with events  Note: Percentages are based on the number of safety population participants in each treatment group. Participants are counted once within each preferred term. Preferred terms are ordered by descending frequency within the vaccine group. | | | | | | | | |
